# EpiInvert, an R application to restore, analyze, compare and forecast epidemiological time series

**DOI:** 10.1101/2024.12.13.24319011

**Authors:** Jean-David Morel, Jean-Michel Morel, Luis Alvarez

## Abstract

The Covid-19 pandemic produced regional time series of incidence, hospital, ICU admission and death. The **EpiInvert** package estimates the incidence trend and the daily reproduction number *R*_*t*_ of any infectious disease, compares related time series such as incidence and death, and provides incidence forecasts. **EpiInvert** is an R package with the following features: (1) *EpiInvert* inputs raw daily incidence time series and the pandemic time serial interval. It outputs a weekly seasonality, an incidence trend and its reproduction number. (2) *EpiIndicators* inputs two related epidemiological time series such as daily incidence and death count. It outputs a daily ratio and delay between both time series. (3) *EpiInvertForecast* inputs an incidence trend obtained by *EpiInvert* and a database of past observed time series. Using the most similar past series, it forecasts the incidence in the next four weeks. **EpiInvert** is in the CRAN repository https://cran.r-project.org/web/packages/EpiInvert/index.html.

## Introduction

Several software are dedicated to the simulation and control of infectious diseases [13, 5, 4], but few analyze epidemiological times series by signal processing techniques as this software does. The underlying three main algorithms have been published in [1, 3, 16, 17] and are now grouped into a single coherent CRAN package [2].

The software inputs epidemiological time series such as incidence, hospital admission, ICU admission or deaths counts and implements four applications:

a. seasonal - trend decomposition [3] of the time series;
b. estimation of the time dependent reproduction number *R*_*t*_ [1] as a component of the above seasonal-trend decomposition. It provides an estimate of *R*_*t*_ several days in advance with respect to the estimate provided by the CRAN package EpiEstim [7];
c. quantitative comparison of related epidemiological time series, estimation of their time shift and ratio [17]. To the best of our knowledge, no other software provides this functionality. This tool estimates, for example, the evolution of the mortality ratio and the time delay between incidence and deaths;
d. short term incidence forecast [16], which proved competitive in the European Covid-19 Forecast Hub.

### EpiInvert: Weekly seasonal - trend decomposition of the daily incidence and calculation of *R*_*t*_

Seasonal - trend decomposition is crucial in time series analysis. One of the standard methods is the STL decomposition [6], [12]. In STL:

1. the seasonal component is allowed to change over time, with user controlled change rate;
2. the smoothness of the trend-cycle also is user-controlled;
3. anomalous values are discarded;

*EpiInvert* also is a seasonal - trend decomposition method, that in addition estimates a daily reproducing number *R*_*t*_ and restores a trend satisfying the classic renewal equation

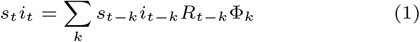

linking the evolution of the daily incidence *i*_*t*_ to the daily case reproduction number *R*_*t*_ [18, 19]. *In (1)*, *s*_*t*_ is the weekly seasonal component and Φ_*k*_ an observed serial interval such as those observed during the Covid-19 pandemic [20, 10, 9]. By introducing *s*_*t*_ in (1) we correct the administrative bias introduced by the way the number of cases is recorded in each country. The epidemiological justification of *EpiInvert* has been published in [1] and [3].

Figure 1 illustrates how *EpiInvert* analyzes a five month long Covid-19 daily incidence in the USA. The first curve is the raw *incidence curve*. The second, *festive bias free incidence* is corrected of the impulses caused by festive days. The third one shows this incidence corrected of the *weekly seasonality bias* caused by almost periodic variations of the administrative measurements. The *seasonality* multiplicative correction factors *s*_*t*_ are shown in the sixth curve of Figure 1. The fourth curve is the fully *restored incidence* (or trend) that satisfies the renewal equation (1). The last curve, *normalized noise* shows the difference between the third *bias free incidence curve* and the *restored incidence*. Statistical tests [3] prove this residual to be uncorrelated noise.

**Fig 1.**
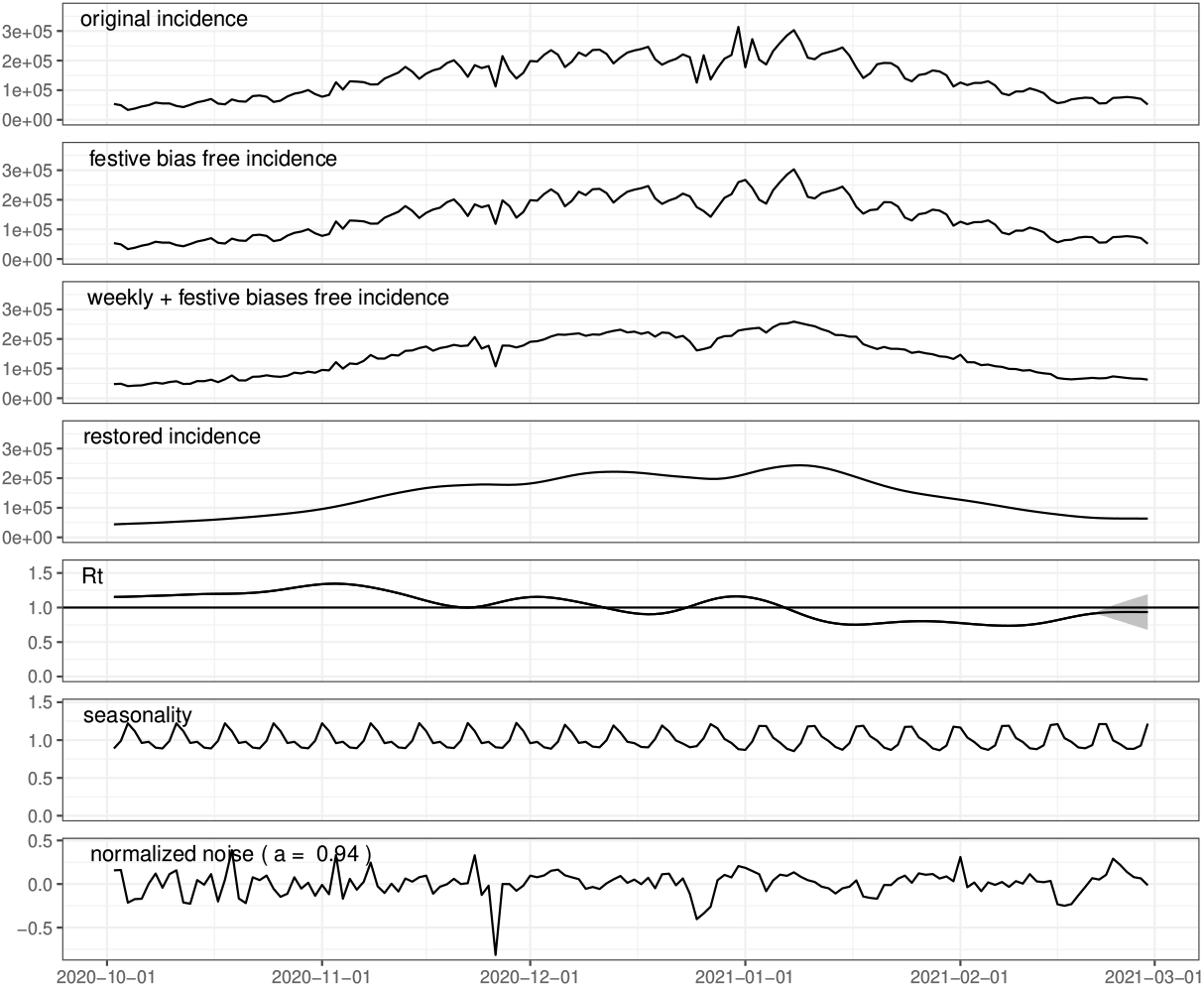
Daily incidence decomposition obtained by **EpiInvert** for the USA using the daily incidence values up to 2021-02-28.

Festive (or anomalous) days, cause sharp oscillation of the registered incidence. The user can signal these days and the algorithm corrects them by redistributing the cases on the anomalous day and its next two days. This does not hinder the analysis of the infectious effects of festive events, as performed in [11] using *EpiInvert*.

*EpiInvert* computes the case reproduction number *R*_*t*_, a key indicator of the epidemic trend, displayed in the fifth curve of Figure 1. When *R*_*t*_ is larger than 1, the pandemic is out of control. When it is smaller, the pandemic loses momentum. The most popular method computing *R*_*t*_ is EpiEstim [8], published as a CRAN R package in [7]. *EpiEstim* [8] uses a simplified version of the renewal equation and removes the weekly seasonal component of the daily incidence by working on a weekly aggregated value. This strategy introduces a several days delay in the estimation of *R*_*t*_, as shown in [1].

### *EpiIndicators* : Establishing the relationship between epidemiological time series

The usual way to establish the relationship between two events in the course of the evolution of an infectious disease, such as infection and death, is to use a cohort of patients and study their evolution. Yet, the number of patients is very small compared to the entire population and the cohort may not correctly represent the population. Furthermore, a cohort is a snapshot of the epidemic situation by the time at its selection time. Therefore, studying the evolution throughout the epidemic of the relation between two epidemiological events requires several successive cohorts. *EpiIndicators* instead uses pairs of public time series to establish a relationship between epidemiological events. The epidemiological time series can be for example the daily or weekly registered incidence, deaths counts, hospitalization admissions and ICU’s admissions. The mathematical foundation and epidemiological verification of this method are published in [17]. The method inputs a pair of time series *f* (*t*) and *g*(*t*) like the ones shown in Figure 2-A. The blue one *f* (*t*) is the Covid-19 incidence in France measured between February 2020 and November 2023 and the red one *g*(*t*) the corresponding death count. The method assumes that both curves are related by a smooth time varying delay, *s*(*t*), such as the delay between cases and deaths. Once the delay between both curves is compensated, the ratio between their values, *r*(*t*), is assumed to be a smooth function interpreted as another pandemic parameter such as the mortality. Therefore, the method computes smooth functions *s*(*t*) and *r*(*t*) such that

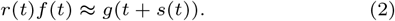

Figure 2-B shows a comparison of *r*(*t*)*f* (*t*) and *g*(*t* + *s*(*t*)) after computing the smooth time shift *s*(*t*) and ratio *r*(*t*) displayed in Figure 2-C. Using an inverse problem technique in signal processing, *EpiIndicators* computes smooth functions *s*(*t*) and *r*(*t*) and *s*(*t*) minimizing the difference between both sides of equation (2). The analyses presented in [17] show how experiments like the one of Figure 2 facilitate the interpretation of the different phases of the pandemic and the progress of sanitary policies in France. They bring a helpful complement to statistical analyses performed on time intervals and more restricted cohorts in [14] and [22].

**Fig 2.**
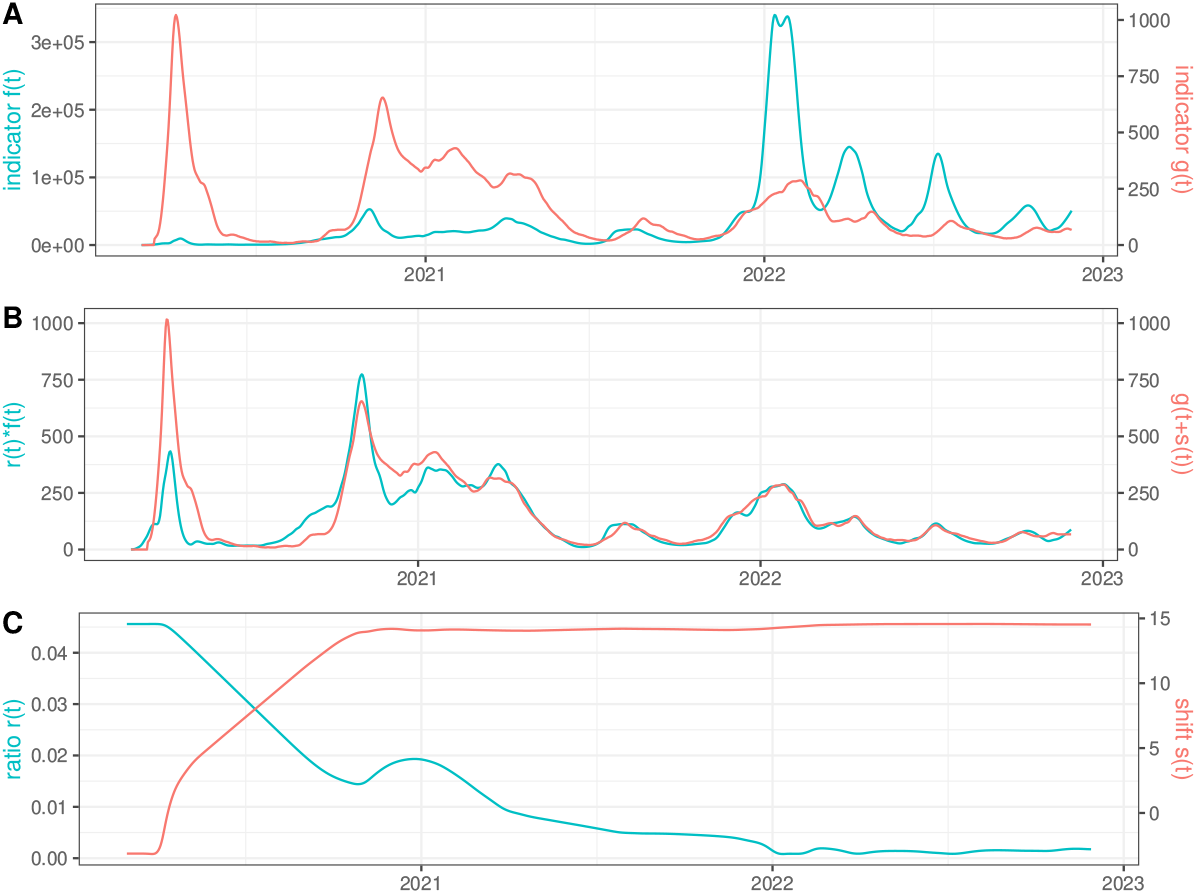
Comparison of the restored incidence, *f* (*t*), and deaths, *g*(*t*), in France, using **EpiIndicators**.

### A learning method for the short time forecast of the daily incidence time series

Classical general purpose forecasting methods such as ARIMA or exponential smoothing [12] can deal with seasonality but the underlying evolution model does not take into account the specifics of an epidemic evolution and the forecasting is only based on the past values of the current incidence curve. *EpiInvertForecast* does not use any model for the epidemic evolution and instead uses a learning procedure based on a large database of incidence time series in many countries. Given a current daily incidence curve observed over the past four weeks, its forecast in the next four weeks is computed by matching it with the four week intervals of all samples in the database, and ranking them by their similarity to the query curve. Then the 28 days forecast is obtained by a statistical estimation combining the values of the next 28 observed days in those similar four week intervals. Using comparison performed by the European Covid-19 Forecast Hub with the current state of the art forecast methods showed that the proposed global learning method, *EpiInvertForecast*, compares favorably to methods forecasting from a single past curve [16]. The method can be seen as an extension of the “method of analogues”, inspired from meteorology and first introduced for epidemiologic forecasting in [21] in predicting influenza activity. Figure 3 shows in green and blue a four weeks forecast of the incidence in the USA.

**Fig 3.**
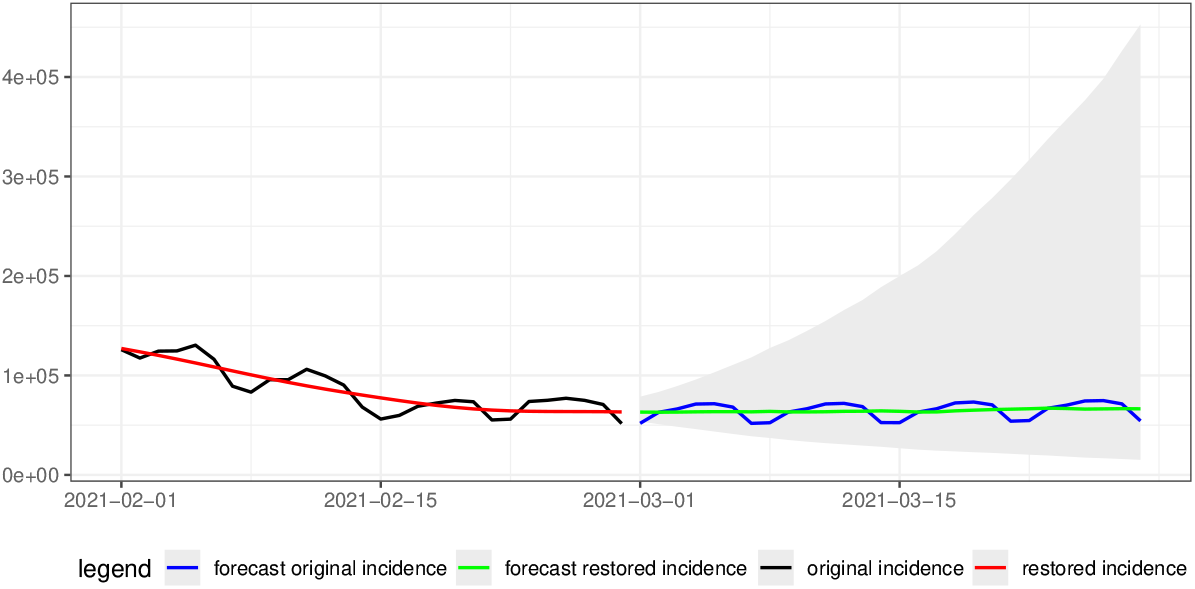
Forecasting of the daily incidence in the USA using **EpiInvertForecast**.

## Implementation

*EpiInvert* inputs the raw incidence *i*_*t*_ and computes the restored incidence *T*_*t*_ or trend, obtained as the second member of the renewal equation (1), and the time varying reproduction number *R*_*t*_.

To perform this inversion, the user disposes of a smoothness parameter limiting variations of the seasonality *s*_*t*_ and of *R*_*t*_. Once *R*_*t*_ and *s*_*t*_ are computed, the trend component *T*_*t*_ (a restored incidence) and the normalized error *ε*_*t*_ are defined by

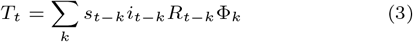

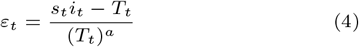

where *a* is computed through an approximation of |*s*_*t*_*i*_*t*_ − *T*_*t*_| by an exponential distribution.

The **EpiInvert** package user interface is implemented in R and published in the R CRAN repository. For computational cost efficiency, the algorithms have been implemented in C++.

The **EpiInvert** package includes the three main functionalities described above, that is *EpiInvert, EpiIndicators* and *EpiInvertForecast* which have associated R functions with the same name.

## Use

The R software package *EpiInvert* is easily installed from the CRAN repository. Once the library is attached, stored data on COVID-19, obtained from the Our World in data organization (https://github.com/owid/covid-19-data/tree/master/public/data), up to 2022-11-28 and official festive days can be loaded for some countries using the R code:

data(owid)

data(festives)

To apply *EpiInvert* to the daily incidence for the USA up to 2021-02-28 use the R code:

sel ← filter(owid,iso code==“USA” & date*<*= “2021-02-28”)

eUSA ← EpiInvert(sel$new cases,”2021-02-28”,festives$USA)

where the first parameter of the *EpiInvert* function call, sel$new cases, is a vector with the daily incidence time series, the second parameter, “2021-02-28”, is the final day of the time series and the third parameter, festives$USA, is a character vector with the festive days in the USA. We visualize in Fig. 1, the daily incidence decomposition obtained by *EpiInvert* using the R code:

EpiInvert plot(eUSA)

the restored incidence corresponds to *T*_*t*_ and the seasonality to the weekly seasonal component *s*_*t*_. *EpiInvert* uses the default serial interval estimated in [15] for COVID-19. It can easily be replaced by the serial interval of another infectious disease.

See *EpiInvertVignette*.*html* in the supplementary material for more details.

Next we use *EpiIndicators* to study the relationship between the restored incidence and deaths in France. To select that data for France, we filter the *owid* dataset using the R code:

sel ← filter(owid,iso code==“FRA”)

Next we build a dataframe with the selection of data that we use :

df ← data.frame(sel$date,

sel$new cases restored EpiInvert,

sel$new deaths restored EpiInvert)

sel$date is a vector with the time series dates,

sel$new cases restored EpiInvert and

sel$new deaths restored EpiInvert, are the restored time series using *EpiInvert* for the incidence, *f* (*t*), and deaths, *g*(*t*), respectively. We apply *EpiIndicators* to these time series using the R code:

FRA ← EpiIndicators (df)

We visualize the results in Fig. 2 using the R code:

EpiIndicators plot (FRA)

See *EpiIndicatorsVignette*.*html* in the supplementary material for more details.

Next, we use *EpiInvertForecast* to perform a short time forecast of the COVID-19 daily incidence in the USA data up to “2021-01-28”. First, load a database of past incidence curves using the R code:

data(restored incidence database)

Then call *EpiInvertForecast* to perform the forecast using the R code:

forecast ← EpiInvertForecast(eUSA,restored incidence database)

where the first parameter of the function call, *eUSA*, is the *EpiInvert* decomposition obtained above for the USA daily incidence data up to 2021-02-28. One can visualize results such as the ones of Fig. 3 using the R code:

EpiInvertForecast plot(res,forecast)

which includes a 95% confidence interval.

See *EpiInvertForecastVignette*.*html* in the supplementary material for more details.

## Discussion

The seasonal-trend decomposition of the daily incidence based on the renewal equation enables a better understanding of the incidence trend, the weekly administrative bias (given by the seasonal component) and the reproduction number *R*_*t*_. Our definition of *R*_*t*_ is based on the renewal equation (1), as showed in [1]. It has the advantage that it reacts sooner to incidence trend variations than other *R*_*t*_ estimation methods.

Establishing the relationship between different epidemiological time series allows analyzing the evolution, throughout the epidemic, of the temporal shift and the ratio between two epidemiological events such as incidence and death. These parameters provide complementary information with respect to the usual cohort analysis.

The proposed daily incidence short time forecasting method using a learning procedure from the past provides a method which is very different from the usual ones. It favorably compares in forecast performance to other methods (see [16]). It therefore is a good candidate to be included in ensemble models. This method requires daily data, but in the case that only weekly aggregated data are available, we can distribute the weekly value equally between the days of the week.

The development of the three presented tools has been carried out in the context of the COVID-19 epidemic, but it can be applied to any infectious disease for which daily or weekly incidence is available. The only required adaptation for each disease is to dispose of an empirical time interval Φ_*t*_.

## Data Availability

All data produced are available online at https://cran.r-project.org/web/packages/EpiInvert/index.html

## Ethics approval

Not applicable

## Data availability

Data publicly available at *Our World in data* (https://covid.ourworldindata.org/data/owid-covid-data.csv)

## Conflit of interest

None declared.

